# Modified-Mindfulness-Based Stress Reduction as a Treatment for Cognitive Recovery in Patients with Minor Stroke: a Randomized Controlled Pilot Study

**DOI:** 10.1101/2024.11.11.24317111

**Authors:** Sophia G. Girgenti, Isabella Dallasta, Erin Lawrence, Dawn Merbach, Jonathan Z. Simon, Rafael Llinas, Neda F. Gould, Elisabeth Breese Marsh

**Affiliations:** Johns Hopkins School of Medicine, Departments of Neurology, Baltimore, MD, USA; Psychiatry and Behavioral Sciences, Baltimore, MD, USA; University of Maryland, Departments of Electrical Engineering, College Park, MD, USA; Biology, College Park, MD, USA

**Author notes:** Corresponding Author: EB Marsh, MD, Johns Hopkins School of Medicine, 600 North Wolfe St, Phipps 446, Baltimore, MD 21287 p., f., e.

**Keywords:** stroke, recovery, mindfulness, function, cognitive networks, cognition

## Abstract

**Background:** Well-developed rehabilitation paradigms exist for post-stroke language and motor impairments. However, no such recovery program has been identified for commonly disabling cognitive deficits in patients following minor stroke. Mindfulness Based Stress Reduction (MBSR) is thought to engage the frontal lobes, improving concentration and attention, and therefore may be an effective option.

**Methods:** We prospectively enrolled a cohort of patients with subacute minor stroke and randomized them to either an 8-week online modified-MBSR course or online traditional Stroke Support Group (SSG). All patients underwent a battery of cognitive tests and measures of patient reported outcomes (PROs) pre- and post-intervention. ANOVA was used to compare changes in scores over time across both groups, as well as a third group of control patients having received neither intervention (n=128).

**Results:** A total of 30 patients were randomized (n=16 for m-MBSR; n=14 for SSG). The average age of the cohort was 65.9 years. Both groups scored similarly on assessments one-month post-stroke and demonstrated increased T-scores on cognitive tasks at the 3-month visit. However, the m-MBSR group showed moderately elevated levels of improvement, specifically in processing speed, executive, and global cognitive function. Level of engagement was not associated with better clinical scores, though was unexpectedly low for both groups.

**Conclusions:** m-MBSR appears to modestly improve frontal lobe activity and demonstrates some success in increasing cognitive performance. However, further studies are needed to determine if it is more efficacious in the chronic stage of recovery when more patients are able to fully engage and actively participate.

## Introduction

Stroke is the fifth leading cause of death in the United States.^1^ With greater than 7.6 million new ischemic strokes globally per year, the estimated cost is over $721 billion US dollars. ^2^ The most common and visible deficits of major strokes, hemiparesis and aphasia, benefit from well-developed evidence-based rehabilitation paradigms.^3–6^ However, the landscape of recovery is changing as acute treatment with intravenous thrombolytics^7^ and mechanical thrombectomy^8,9^ have allowed larger and more disabling strokes to be treated early, decreasing infarct size and resulting in a greater number of “minor” strokes.^10^ While minor strokes typically spare patients from dependency on long-term care and often allow them to maintain overall independence, they can still be debilitating, resulting in cognitive dysfunction, particularly with respect to executive function, attention, and decision making.^11–13^ These deficits can be observed on evaluations such as the Montreal Cognitive Assessment (MoCA) and appear consistent across the majority of patients, regardless of stroke size and location.^13^ Patients with minor stroke are often unable to return to work due to these cognitive deficits, significantly impacting quality of life and making depression and anxiety common. Unfortunately, apart from the Stroke Support Groups typically offered at hospitals to give patients and their families an outlet to discuss these difficulties with professionals, treatment and rehabilitation options - specifically for cognitive dysfunction - are significantly lacking.

In search of a novel intervention to address cognitive symptoms associated with minor stroke, mindfulness-based stress reduction (MBSR) is an encouraging alternative. MBSR is a group-based intervention combining meditation, yoga, and body awareness to help individuals cope with stress and better handle life’s challenges. Mindfulness cultivates a purposeful, non-judgmental awareness and acceptance of the present mindset. Originating from the ancient practices of Eastern Buddhist monks, this practice is believed to engage the attention networks and frontal lobes, with neuroimaging studies showing activation of frontal cortex as well as changes in frontal activity with meditation.^14–16^ These areas are known to play a significant role in executive function, attention, and decision making - all activities hindered in patients with cognitive dysfunction following minor stroke. Mindfulness training in general has been shown to improve health, quality of life, social functioning, and mental health outcomes measured before and after the intervention.^17,18^ This makes it an appealing therapeutic option for patients experiencing depression, anxiety, and stress, all common symptoms in survivors of minor stroke.^19^ MBSR has shown potential benefit in a heterogenous population of patients, with conditions ranging from traumatic brain injury to cancer, depression, and diabetes.^20–29^ Improvements have also been shown in patients with chronic stroke.^30^ However, no existing studies have investigated the effects of MBSR on stroke in the early stages of recovery when patients show the most improvement, and more specifically, on the growing population of patients with minor strokes who exhibit difficulties with attention and focus.

Patients experiencing minor stroke have tremendous potential to return to their prior level of function; however, many fail to successfully re-integrate into society.^10,19^ Counseling on eventual improvement of symptoms can be helpful, but, understandably, many desire a treatment that will lead to a faster and fuller recovery. MBSR provides a novel, non-pharmacologic approach that may be effective by targeting areas of abnormality implicated in neuroimaging studies such as the frontoparietal cortex.^31^ In this study, we explore the effectiveness of MBSR on improving patient-reported outcomes and cognition in patients with recent minor stroke.

## Methods

This was a prospective, randomized, longitudinal pilot study spanning two years. Patients admitted for acute stroke to our tertiary referral center were scheduled for a follow-up appointment at the Johns Hopkins Bayview Stroke Intervention Clinic (BaSIC) approximately 6 weeks (+/- 2 weeks) post-infarct. Their data were entered into our Stroke Registry, a HIPAA-compliant, IRB-approved database. Participants were screened before presenting for their first follow-up appointment after hospital discharge and those meeting inclusion criteria were then recruited, consented, randomized, and tested at their clinic visit. After the 8-week intervention, individuals were seen again for repeat evaluation. The study was approved by the Johns Hopkins Institutional Review Board and all participants provided written informed consent at the time of enrollment.

### Inclusion and Exclusion Criteria

All patients recruited were adults (≥18 years) presenting with their first clinical ischemic stroke. In order to capture small deep lacunes that presented with motor deficits during the acute period, we defined “minor stroke” as a small area of infarction resulting in an admission NIH Stroke Scale (NIHSS) score of 10 or less. This avoided the inclusion of larger cortical lesions localizing to areas that typically cause disrupted cognition, significant hemiparesis, aphasia, or hemispatial neglect. All participants demonstrated evidence on brain imaging of acute ischemic stroke. Strokes were confirmed and infarct volumes were determined using diffusion-weighted magnetic resonance imaging (MRI) and the lesion tool available on the CareStream platform. Lesions were unilateral and supratentorial, without large vessel involvement. Patients had good premorbid baseline function (modified Rankin Scale [mRS] score ≤2).

Patients were excluded if they presented with intracranial hemorrhage, a multifocal stroke involving multiple vascular distributions, or evidence of aphasia or neglect on examination. Those without an MRI or who had no abnormality on diffusion-weighted imaging (consistent with a transient ischemic attack (TIA) or diffusion-negative stroke) were also excluded. Non– native English speakers were excluded, as well as those with a history of significant dementia, prior clinical stroke, neurologic disease, untreated hearing loss, or psychiatric illness. Other exclusion criteria included inability to attend weekly sessions of MBSR or Stroke Support Group (SSG) and inability to return to clinic following intervention. If the patient had another stroke at any point prior to completion of the study, they were excluded from further participation. For those lost-to-follow-up, data was censored at the time they were lost.

### Interventions

At the time of enrollment, participants were randomized to either 8 weeks of an online modified-Mindfulness-Based Stress Reduction (m-MBSR) program or an online conventional Stroke Support Group (SSG). Though traditionally designed as in-person programs, interventions were converted to an online platform due to the COVID pandemic. All participants were provided with an iPad with the Zoom application installed to connect to the assigned weekly meeting. Individuals without home high-speed internet were also provided with a Hotspot in order to ensure connectivity. If necessary, participants were taught how to use the iPad and Zoom application by a member of the study team prior to the first meeting and the study coordinator was on the call for every group meeting to assist with any technological difficulties.

The MBSR program was run by a board-certified psychologist experienced in MBSR administration. Treatment cohorts were assembled at the beginning of each month with patients recruited the month prior, resulting in groups varying in size from 5 to 15 participants. As the sessions took place online, the MBSR program administered was a modified version (m-MBSR). Participants met once a week for 1.5 hours over Zoom, rather than the usual 2.5 hours in person, and the silent retreat at week 7 was 3 hours instead of the traditional 7. In place of the full yoga routine typically administered in person, the modified version also used a gentle, seated stretching to utilize similar calming body movements. In addition to the weekly meeting, patients were provided with meditation resources and guided audio to practice the concepts each week as a part of the program.

To control for socialization as a confounding factor, an online Stroke Support Group (SSG) was used as the comparator intervention. This SSG program was run by the Stroke Program’s nurse practitioners and was also modified due to the pandemic, meeting once a week for 40 minutes over Zoom rather than once a month for 2 hours in person. Topics pertaining to stroke recovery, the effect stroke has on patients and their caretakers, and steps patients can take to prioritize their health were discussed. Patients were encouraged to actively participate with questions and comments.

For additional comparison, an existing study population of 128 age-similar patients with minor stroke who did not undergo either intervention but had cognitive testing at similar time points was also included in the study as a control arm to evaluate the hypothesis that both m-MBSR and online SSG may be more effective than no intervention on improving patient reported outcomes and cognitive measures. These patients were part of our existing stroke registry, and evaluated at 1- and 6-months post-stroke as part of routine clinical care.

### Engagement

Engagement was assessed over the course of participation based on the hypothesis that in order to achieve clinical and radiographic benefits one must actively participate in therapy. Engagement was defined as the level of alertness displayed by the participant throughout the class, attention paid to the screen during presentations, ability to follow directions, participation, and the amount of homework completed if assigned. Based on this definition, the leaders of the interventions and the study coordinator observed each patients’ participation level including eye contact and whether they responded to the presenter’s questions and prompts or asked their own questions. It was also noted whether they were completing other tasks throughout the session. Initially, engagement was assessed for each session; however clear patterns emerged allowing for creation of a single variable-“engaged” or “not engaged”-representative of participation over the entire intervention period. Any disagreements between reviewers (n=2) were resolved through consensus.

### Clinical and Cognitive Testing

Patient-reported outcome metrics evaluating overall function, mood, and satisfaction were assessed pre- and post-intervention, along with cognitive measures. To evaluate patient-reported outcomes, participants were administered the Stroke Impact Scale (function), Patient Health Questionnaire (depression), Functional Assessment of Chronic Illness Therapy (FACIT) (fatigue), Barthel Index for Activities of Daily Living (function), and Patient Reported Outcomes Measurement Information System (PROMIS) measures (mood, fatigue, and ADLs) at each clinic visit.

A battery of cognitive tests, developed in conjunction with our neuropsychologist to be efficient yet sensitive to cognitive dysfunction commonly faced by those with minor stroke,^13^ was used to evaluate verbal and spatial memory, motor and processing speed, and executive function. Tests included: the Delis-Kaplan Executive Function System (D-KEFS),^32^ Hopkins Verbal Learning Test,^33^ Brief Visuospatial Memory Test–Revised,^34^ Symbol Digit Modalities Test,^35^ and Grooved Pegboard Test.^36^ The Montreal Cognitive Assessment (MoCA)^37^ was also included as a brief screen of global cognition. T-scores were used to describe normative data. If a given task was traditionally described using a z-score (cutoff −4 to 4), it was converted to a *T*-score for consistency and to allow averaging of scores across tasks. Additional information, including patient demographics (age, sex, self-identified race, and education), stroke characteristics (admission and discharge NIHSS ^38^ score, infarcted hemisphere, lesion volume, and cortical *vs.* subcortical location), social support (living with someone at home), functional baseline (pre-stroke mRS score), and medical comorbidities (history of smoking, hypertension, diabetes, depression, and Charlson Comorbidity Index), was also collected.

### Cognitive Analysis

For each cognitive test, *T*-scores were calculated from raw scores using the age-specific normative data according to the corresponding test manual. Cognitive tests were then divided into 6 different domains. Within each domain, *T-*scores were averaged to generate a composite domain score: *Verbal Memory* - Hopkins Verbal Learning Test total learning and Hopkins Verbal Learning Test delayed recall; *Motor Processing Speed -* D-KEFS Trail Making trial 5, Grooved Pegboard Test dominant hand, and Grooved Pegboard Test nondominant hand; *Spatial Memory -* Brief Visuospatial Memory Test–Revised total learning and Brief Visuospatial Memory Test–Revised delayed recall; *Processing Speed -* Symbol Digit Modalities Test written trial, Symbol Digit Modalities Test oral trial, and D-KEFS Trail Making trial 1; *Executive Function -* D-KEFS letter fluency, D-KEFS category fluency, and D-KEFS Trails Making trials 2, 3, and 4; and *Overall Global Function*.

### Statistical Analysis

Data were analyzed using Stata version 14 (College Station, TX). To determine the impact of m-MBSR on post-stroke cognitive impairment, the primary aim of this study, ANOVA was used to compare performance on PROs and cognitive tasks between all 3 groups at each time point, as well as averaged cognitive domains. Changes in score over time were also compared. Based on a sample size of 15 patients per group, we anticipated 80% power to detect a difference in mean performance of 0.53 points between groups, a relatively mild difference in outcome.

To assess the impact of engagement on outcome, independent t-tests were used within the m-MBSR cohort to compare performance for those engaged versus not engaged.

## Results

Thirty patients were randomized to Stroke Support Group (n=14, 7 male (50%)) or modified-Mindfulness Based Stress Reduction (n=16, 9 male (56%)). Repeat evaluation was performed following the 8-week intervention. By the second clinic visit, one patient had dropped out of the study and a second was censored due to recurrent stroke resulting in 3-month clinical data from 28 patients (SSG n=14, m-MBSR n=14). Seventy-nine (62%) of the 128 patients in the control group who were evaluated at one month but did not undergo intervention returned for follow-up testing. Their follow-up was somewhat later than the other two groups, on average 6.8 (compared to 4.1) months post-stroke.

### Group Comparisons - Demographics and Stroke Severity

The average age of the entire cohort was 64.7 years. Forty-nine percent was male; 27% black. The three groups were similar in age, race, and sex. Medical comorbidities, measured by the Charleston Comorbidity Index (CCI) and premorbid mRS scores were consistent across intervention groups (SSG: 1.8 and 0.31 respectively; m-MBSR: 1.4 and 0.31); though pre-stroke mRS was significantly lower for the much larger control group (0.06, p=0.004) (see Table 1 for full details). Stroke severity was comparable across all groups, with a mean admission and discharge NIHSS for the entire cohort of 2.7 and 1.6 respectively. The infarct volumes were small, ranging from 0.06 to 18.9 cc, with greater than 85% of recruited patients having an infarct size under 5cc. Ten participants (71.4%) in SSG and 10 in Mindfulness (62.5%) had infarcts within the left hemisphere – a slightly higher proportion than controls (n=60 (41.2%)). While the percentage of patients with isolated subcortical strokes was not significantly different across groups (SSG: 8 (57.1%), m-MBSR: 10 (62.5%), control: 74 (59.2%), p=0.953), the percentage of patients with isolated cortical locations did differ (SSG: 6 (42.9%), m-MBSR: 6 (37.5%), control: 19 (15.2%), p=0.009).

**Table 1.**
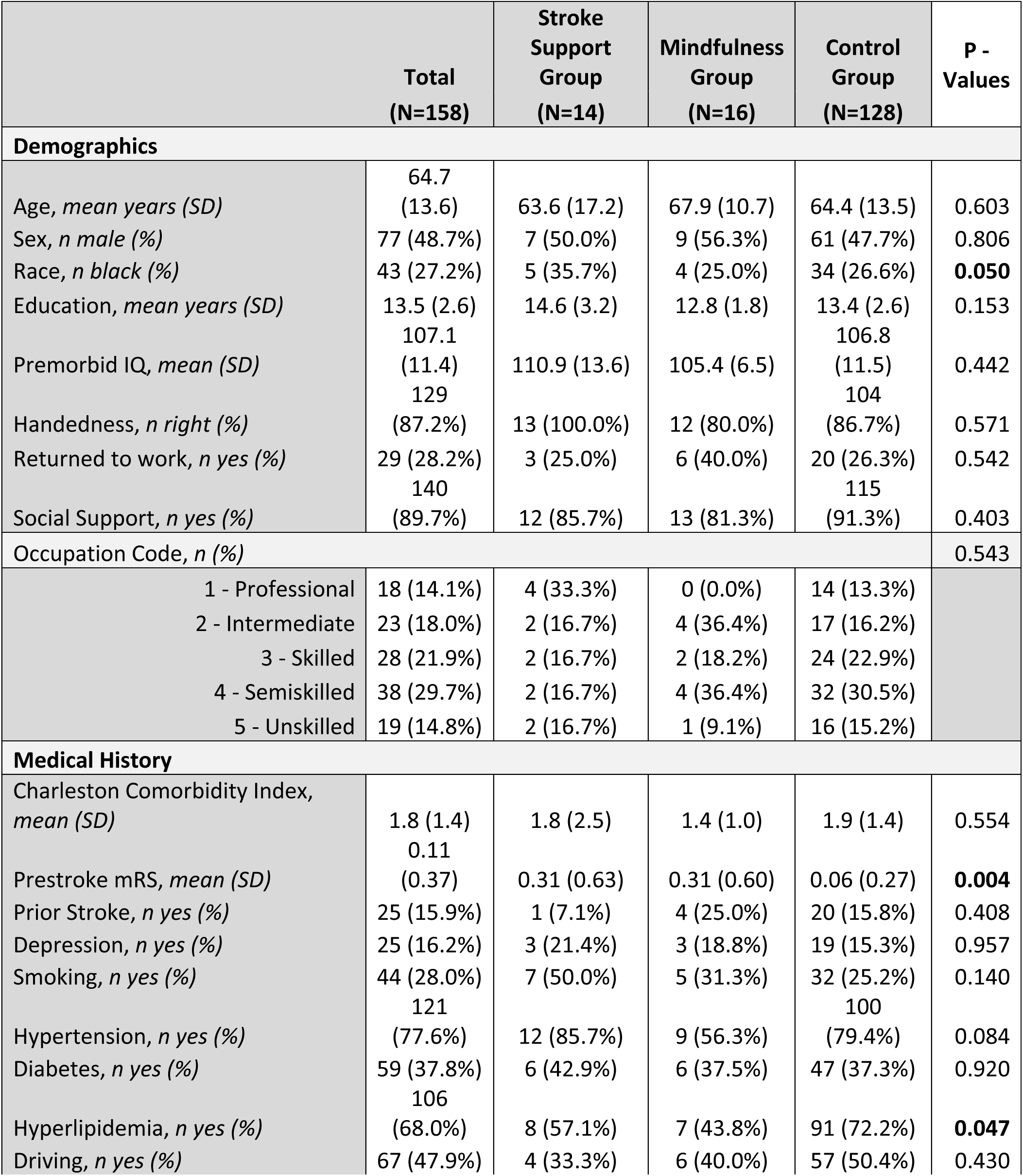
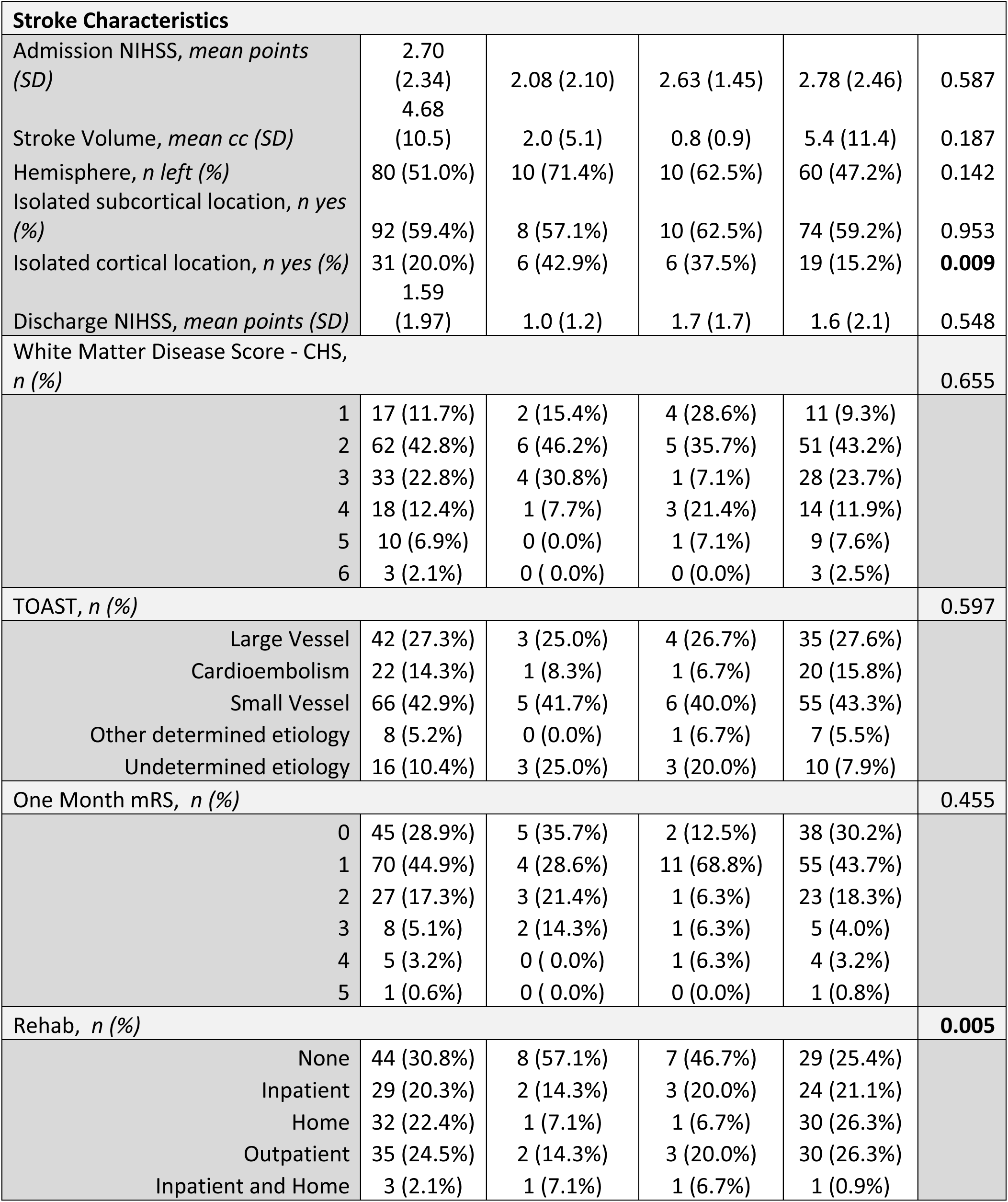
Patient Characteristics.

### Group Comparisons – Functional Assessment and Patient Reported Outcomes

Group comparisons of cognitive function and PROs are detailed in Table 2. Functional outcomes at the baseline visit were similar across groups. Those randomized to online SSG had an average NIHSS of 1.0 and mRS of 1.1, compared to an NIHSS of 1.6 and mRS of 1.3 in the m-MBSR group (p=ns). All groups saw improvement by their post-intervention visit, as documented in Table 1.

**Table 2.** Group Performance.

|  | Baseline |  |  |  | Follow-up |  |  |  |
| --- | --- | --- | --- | --- | --- | --- | --- | --- |
|  | Stroke Support Group (N=14) | Mindfulness Group (N=16) | Control Group (N=128) | P-value | Stroke Support Group (N=14) | Mindfulness Group (N=14) | Control Group (N=79) | P-value |
|  | **Stroke Characteristics, mean (SD)** |  |  |  |  |  |  |  |
| Barthel Index (BI) | 95.0 (10.2) | 96.3 (10.4) | 97.4 (8.7) | 0.587 | 98.3 (5.8) | 100.0 (0.0) | 99.3 (3.2) | 0.509 |
| NIH Stroke Scale (NIHSS) | 1.00 (1.57) | 1.63 (1.89) | 0.82 (1.48) | 0.141 | 0.58 (1.00) | 0.40 (0.52) | 0.45 (1.23) | 0.917 |
| Modified Rankin Scale (mRS) | 1.14 (1.10) | 1.25 (1.00) | 1.09 (1.03) | 0.831 | 1.17 (1.27) | 0.90 (0.74) | 0.72 (0.90) | 0.299 |
|  | **Patient-Reported Outcomes, mean (SD)** |  |  |  |  |  |  |  |
| FACIT | 36.4 (8.7) | 32.0 (14.9) | 37.3 (11.7) | 0.374 | 37.6 (10.4) | 30.7 (13.0) | 38.2 (11.2) | 0.164 |
| PHQ-9 | 5.2 (5.9) | 6.7 (7.3) | 4.9 (5.8) | 0.655 | 4.9 (3.9) | 6.1 (6.9) | 4.4 (5.2) | 0.670 |
| Likert-Symptoms | 6.0 (1.3) | 6.2 (1.2) | 5.5 (1.6) | 0.312 | 5.4 (1.3) | 5.3 (1.9) | 5.7 (1.5) | 0.771 |
| Likert-QOL | 6.1 (1.1) | 5.7 (2.2) | 5.2 (1.9) | 0.349 | 5.1 (1.8) | 5.2 (1.9) | 5.5 (1.6) | 0.779 |
| Percent Recovered | 79.8 (18.2) | 76.8 (24.1) | 76.3 (23.3) | 0.918 | 76.4 (19.3) | 77.5 (19.3) | 80.6 (17.6) | 0.778 |
| **Stroke Impact Scale** |  |  |  |  |  |  |  |  |
| Strength | 90.8 (11.1) | 72.5 (23.1) | 79.1 (22.1) | 0.294 | 76.7 (22.5) | 71.7 (24.8) | 80.2 (20.2) | 0.618 |
| Memory and Thinking | 86.2 (13.4) | 77.1 (19.6) | 87.9 (16.7) | 0.224 | 75.7 (12.9) | 84.3 (11.5) | 88.1 (12.9) | 0.086 |
| Mood | 86.6 (11.7) | 78.5 (9.0) | 83.9 (17.2) | 0.583 | 76.4 (9.8) | 74.2 (22.5) | 83.9 (13.8) | 0.233 |
| Communication | 96.2 (6.9) | 89.2 (13.7) | 92.3 (15.8) | 0.687 | 81.1 (20.2) | 90.2 (15.9) | 93.5 (10.9) | 0.112 |
| Activities of Daily Living | 98.7 (1.6) | 85.0 (22.0) | 89.2 (18.8) | 0.383 | 78.0 (28.3) | 86.0 (15.2) | 95.6 (12.2) | **0.025** |
| Mobility | 96.3 (5.4) | 89.2 (19.6) | 85.0 (19.3) | 0.337 | 77.8 (20.7) | 81.8 (19.7) | 90.7 (13.1) | 0.102 |
| Fine Motor Movements | 96.0 (8.0) | 91.4 (21.0) | 84.2 (23.4) | 0.367 | 68.8 (35.6) | 83.0 (21.8) | 91.6 (12.6) | 0.015 |
| Socialization | 92.5 (6.9) | 86.8 (27.4) | 78.5 (21.5) | 0.217 | 85.6 (10.1) | 75.5 (33.2) | 86.7 (17.0) | 0.450 |
| **PROMIS Measures** |  |  |  |  |  |  |  |  |
| Anxiety (lower better) | 8.6 (0.9) | 6.3 (0.9) |  | 0.092 | 7.3 (1.1) | 7.1 (0.9) |  | 0.868 |
| Depression (lower better) | 6.4 (0.9) | 6.1 (1.1) |  | 0.872 | 5.7 (0.8) | 5.9 (0.9) |  | 0.841 |
| Fatigue (lower better) | 9.5 (1.0) | 10.6 (1.3) |  | 0.531 | 7.6 (0.8) | 9.9 (1.1) |  | 0.114 |
| Sleep (lower better) | 10.4 (1.1) | 9.1 (1.0) |  | 0.351 | 10.5 (0.9) | 9.0 (1.1) |  | 0.302 |
| Social (higher better) | 14.6 (1.4) | 16.8 (1.2) |  | 0.259 | 17.1 (1.2) | 15.3 (1.2) |  | 0.302 |
| Pain (lower better) | 6.9 (1.2) | 7.8 (1.4) |  | 0.638 | 5.8 (0.9) | 9.0 (1.6) |  | 0.103 |
| Intensity (lower better) | 2.1 (0.8) | 2.6 (0.8) |  | 0.652 | 1.8 (0.8) | 4.0 (0.9) |  | 0.092 |
| Physical (higher better) | 16.1 (1.4) | 15.9 (1.2) |  | 0.887 | 17.3 (1.1) | 15.6 (1.3) |  | 0.350 |
| Anxiety 8a (lower better) | 15.7 (1.6) | 13.5 (1.9) |  | 0.377 | 15.3 (1.8) | 14.7 (2.2) |  | 0.835 |
| Depression 8a (lower better) | 13.1 (1.7) | 12.3 (1.6) |  | 0.725 | 11.2 (1.3) | 12.4 (1.8) |  | 0.594 |
| Self Efficacy 4a (higher better) | 14.9 (1.4) | 16.2 (0.9) |  | 0.446 | 15.5 (1.0) | 14.5 (1.5) |  | 0.579 |
| **Cognitive Testing T Scores, mean (SD)** |  |  |  |  |  |  |  |  |
| MoCA | 22.5 (5.9) | 23.5 (5.2) | 24.3 (3.7) | 0.231 | 23.7 (6.0) | 25.5 (3.7) | 25.5 (3.4) | 0.276 |
| **Peg Board Task (TIMES)** |  |  |  |  |  |  |  |  |
| Dominant Hand | 23.7 (67.1) | 14.8 (61.2) | 22.1 (65.8) | 0.595 | 21.7 (67.4) | 21.9 (66.5) | 27.5 (71.0) | 0.578 |
| Nondominant Hand | 25.8 (68.5) | 20.6 (66.9) | 22.1 (66.1) | 0.433 | 23.1 (65.9) | 22.3 (67.7) | 26.8 (67.9) | 0.979 |
| **DKEFS Letter Fluency Task** |  |  |  |  |  |  |  |  |
| Letter Fluency | 43.1 (16.0) | 42.3 (14.4) | 46.9 (13.8) | 0.345 | 44.7 (14.4) | 50.4 (13.7) | 49.5 (13.4) | 0.445 |
| **DKEFS Category Fluency Task** |  |  |  |  |  |  |  |  |
| Category Fluency | 39.9 (13.1) | 43.8 (10.3) | 44.7 (13.0) | 0.401 | 44.4 (16.2) | 46.6 (9.8) | 46.8 (13.3) | 0.820 |
| **DKEFS Category Switching Task** |  |  |  |  |  |  |  |  |
| Category Switching | 43.6 (17.0) | 45.6 (9.1) | 45.4 (13.1) | 0.889 | 52.4 (14.9) | 51.2 (13.8) | 47.4 (12.3) | 0.287 |
| Switching Accuracy | 45.7 (16.0) | 48.8 (8.6) | 46.1 (12.0) | 0.684 | 53.1 (13.7) | 50.2 (13.3) | 47.5 (11.8) | 0.250 |
| **DKEFS Trails Task** |  |  |  |  |  |  |  |  |
| Trail 1 | 33.8 (14.3) | 35.4 (14.1) | 30.6 (13.5) | 0.330 | 36.6 (16.6) | 40.1 (12.2) | 44.7 (13.6) | 0.097 |
| Trail 2 | 38.8 (13.2) | 43.5 (16.2) | 41.0 (13.3) | 0.654 | 36.3 (14.4) | 44.8 (12.6) | 45.5 (13.2) | 0.061 |
| Trail 3 | 36.0 (12.7) | 39.8 (15.7) | 41.5 (13.2) | 0.352 | 39.1 (13.8) | 43.1 (14.9) | 46.1 (11.5) | 0.132 |
| Trail 4 | 42.7 (11.8) | 41.6 (14.6) | 47.0 (13.5) | 0.217 | 42.6 (15.1) | 49.6 (13.5) | 48.2 (13.0) | 0.323 |
| Trail 5 | 33.7 (12.4) | 43.1 (9.6) | 44.3 (11.5) | **0.008** | 34.3 (14.2) | 41.3 (13.5) | 45.8 (11.2) | **0.004** |
| **HVLT task** |  |  |  |  |  |  |  |  |
| Total Recall | 32.9<br>(7.7) | 33.1 (16.4) | 30.5<br>(9.0) | 0.460 | 36.9<br>(12.5) | 32.0 (12.0) | 33.5<br>(9.6) | 0.431 |
| Delayed Recall | 33.9<br>(11.8) | 30.8 (11.7) | 30.0<br>(10.1) | 0.414 | 34.9<br>(13.4) | 32.6 (11.0) | 34.0<br>(11.1) | 0.871 |
| Recognition | 40.7<br>(14.1) | 35.1 (10.6) | 33.3<br>(12.9) | 0.119 | 37.6<br>(14.5) | 38.0 (10.5) | 35.7<br>(11.8) | 0.734 |
| Retention | 38.4<br>(14.0) | 37.5 (19.9) | 35.6<br>(14.6) | 0.748 | 36.4<br>(14.3) | 36.9 (14.9) | 39.1<br>(14.3) | 0.732 |
| **BVMT Task** |  |  |  |  |  |  |  |  |
| Total Recall | 34.3<br>(12.6) | 44.1 (14.2) | 41.6<br>(12.6) | 0.081 | 40.2<br>(15.1) | 42.8 (16.2) | 43.7<br>(14.3) | 0.712 |
| Delayed Recall | 39.1<br>(13.1) | 46.9 (13.9) | 42.7<br>(13.8) | 0.312 | 37.6<br>(16.5) | 46.3 (17.6) | 44.7<br>(15.1) | 0.256 |
| **SDMT** |  |  |  |  |  |  |  |  |
| Written | 41.4<br>(14.3) | 37.6 (11.3) | 41.5<br>(12.7) | 0.508 | 42.0<br>(13.2) | 43.8 (13.2) | 45.0<br>(12.1) | 0.697 |
| Oral | 40.5<br>(12.4) | 40.1 (12.7) | 37.9<br>(11.9) | 0.623 | 42.6<br>(14.1) | 42.9 (12.4) | 39.9<br>(11.5) | 0.572 |

Baseline Patient Reported Outcomes (PROs) were also similar across groups. While the modified-MBSR group displayed slightly higher levels of depression on the PHQ-9 (6.7 versus 5.2 (p=ns)), the online SSG reported higher anxiety levels (8.6 versus 6.3 (p=ns)) on the PROMIS anxiety section. Individuals in the SSG reported slightly higher scores on the Stroke Impact Scale for every category, indicating higher mood, better mobility, more socialization and ADLs, and feeling less cognitively slow although results were not statistically significant. Interestingly, by the second visit, the higher Stroke Impact Scale scores for 5 of the 8 categories switched to the m-MBSR group, though remained non-significant. The control group did report significantly better scores for the Activities of Daily Living (ADLs) and Fine Motor categories compared to those in the SSG at post-intervention follow-up, which may have been secondary to the extended period of time allowed for recovery. However, there were no other major differences.

### Group Comparisons - Cognitive Testing

Cognitive testing scores were also similar at the baseline time point between groups, with average (SD) MoCA scores of 22.5 (5.9), 23.5 (5.2) and 24.3 (2.7) for the SSG, m-MBSR, and control groups, respectively. While overall scores remained similar at the second visit, the m-MBSR average (SD) MoCA score of 25.5 (3.7) appeared more consistent with the control group’s 25.5 (3.4) than the SSG, with an average of only 23.7 (6.0). As expected, cognitive scores improved consistently for all groups between the first and second visits, though insignificant. The SSG group demonstrated a slower D-KEFS Trails Task 5 score at baseline (T = 33.7 (12.4)), almost one standard deviation below the m-MBSR group and a full SD below the control group (T = 43.1 (9.6) and T = 44.3 (11.5) respectively, p=0.008). This difference persisted at follow-up (p=0.004). There were no other significant differences in performance across groups.

### Changes in Scores Post-Intervention

Given that all groups were expected to recover and that differences were modest between intervention groups at both time points, a heat map was generated to better visualize the pattern of differences in improvement over time between groups (Figure 1). Those participating in m-MBSR showed greater degrees of improvement in objective functional measures, such as the BI and mRS, and patient reported assessments of function (SIS, Likert scales perception of recovery), while the SSG showed greater degrees of improvement on the patient-reported PROMIS measures evaluating emotional well-being. The SSG also improved to a greater extent in the areas of verbal and spatial memory, while m-MBSR participants demonstrated more improvement in processing speed, executive function, and global cognition.

**Figure 1.**
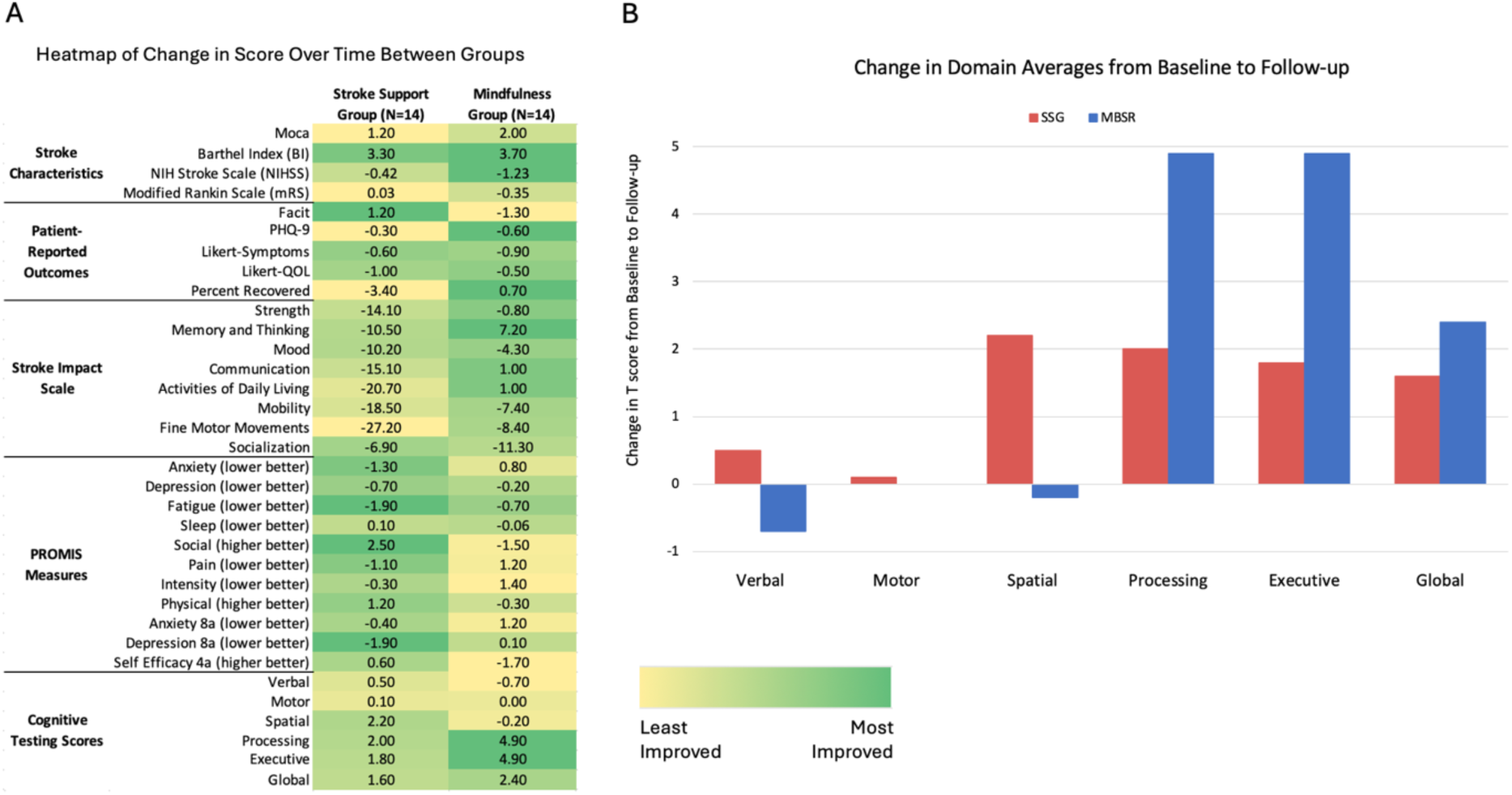
Group Differences. a) Patients in the m-MBSR group demonstrated a greater degree of change over time in objectively measured outcomes of performance following intervention, while the SSG tended to show more improvement in patient-reported metrics. b) Specifically, patients in the m-MBSR group demonstrated more improvement than SSG in processing speed, executive function, and global processing domains

### Engagement

Nine individuals (60%) within the m-MBSR group were actively engaged throughout the duration of the intervention, while 6 were not. Eleven of 14 were engaged during the SSG (78.6%). Within the m-MBSR group, we saw no significant differences in cognitive performance between the engaged and unengaged groups (Table 3). However, it was notable that the engaged group tended to perform better both pre- and post-intervention compared to the unengaged group.

**Table 3.** Effect of Engagement.

|  | Baseline |  |  | Follow-up |  |  |
| --- | --- | --- | --- | --- | --- | --- |
|  | Unengaged<br>(n=6) | Engaged<br>(n=9) | P-value | Unengaged<br>(n=5) | Engaged<br>(n=9) | P-value |
| **Stroke Characteristics, mean (SD)** |  |  |  |  |  |  |
| MoCA | 21.3 (2.2) | 25.1 (1.7) | 0.192 | 24.4 (2.3) | 26.1 (0.9) | 0.428 |
| Barthel Index (BI) | 95.8 (4.2) | 96.1 (3.9) | 0.963 | 100 (0.0) | 100 (0.0) | 1.000 |
| NIH Stroke Scale (NIHSS) | 1.67 (1.09) | 1.67<br>(0.50) | 1.000 | 0.2 (0.2) | 0.6 (0.2) | 0.242 |
| Modified Rankin Scale<br>(mRS) | 1.17 (0.40) | 1.33<br>(0.37) | 0.772 | 0.6 (0.2) | 1.2 (0.4) | 0.217 |
| **Patient-Reported Outcomes, mean (SD)** |  |  |  |  |  |  |
| FACIT | 34.8 (7.1) | 28.6 (5.6) | 0.519 | 31.5 (6.9) | 30.2 (5.6) | 0.884 |
| PHQ-9 | 6.2 (4.2) | 7.2 (2.4) | 0.842 | 8.5 (4.8) | 4.5 (2.0) | 0.401 |
| Likert-Symptoms | 6.5 (0.5) | 6.0 (0.6) | 0.571 | 5.0 (1.2) | 5.5 (0.8) | 0.732 |
| Likert-QOL | 5.6 (0.9) | 5.8 (1.2) | 0.896 | 5.0 (1.0) | 5.3 (0.8) | 0.818 |
| Percent Recovered | 72.0 (11.6) | 80.8 (9.9) | 0.573 | 85.0 (2.9) | 72.5 (9.8) | 0.346 |
| **Stroke Impact Scale** |  |  |  |  |  |  |
| Strength | 80.0 (10.8) | 65.0<br>(12.6) | 0.401 | 77.5 (13.6) | 60.0<br>(15.0) | 0.478 |
| Memory and Thinking | 85.7 (9.5) | 68.6 (9.3) | 0.243 | 83.5 (7.0) | 85.7 (5.7) | 0.855 |
| Mood | 76.1 (5.2) | 80.5 (3.7) | 0.507 | 60.7 (10.4) | 94.4 (3.3) | 0.090 |
| Communication | 88.6 (6.7) | 89.7 (6.9) | 0.910 | 83.4 (16.7) | 97.1 (2.9) | 0.500 |
| Activities of Daily Living | 87.5 (11.8) | 82.5<br>(11.7) | 0.774 | 90.0 (10.0) | 80.0<br>(10.0) | 0.550 |
| Mobility | 99.3 (0.7) | 81.7<br>(12.1) | 0.275 | 87.4 (12.6) | 73.4<br>(13.4) | 0.515 |
| Fine Motor Movements | 100.0 (0.0) | 85.0<br>(13.7) | 0.397 | 76.0 (24.0) | 90.0 (6.0) | 0.629 |
| Socialization | 98.3 (1.7) | 78.1<br>(17.7) | 0.381 | 60.0 (20.8) | 98.8 (1.3) | 0.245 |
| **PROMIS Measures** |  |  |  |  |  |  |
| Anxiety (lower better) | 7.5 (2.3) | 5.7 (0.7) | 0.391 | 8.6 (2.3) | 6.2 (0.7) | 0.240 |
| Depression (lower<br>better) | 8.5 (2.6) | 4.8 (0.4) | 0.108 | 6.8 (2.3) | 5.4 (0.6) | 0.479 |
| Fatigue (lower better) | 8.7 (2.4) | 12.3 (1.6) | 0.203 | 8.2 (2.2) | 10.9 (1.2) | 0.265 |
| Sleep (lower better) | 10.7 (2.0) | 8.4 (1.0) | 0.292 | 9.0 (2.4) | 9.0 (1.3) | 1.000 |
| Social (higher better) | 18.3 (1.7) | 15.3 (1.8) | 0.269 | 16.8 (2.2) | 14.4 (1.5) | 0.378 |
| Pain (lower better) | 6.2 (2.0) | 9.2 (2.1) | 0.327 | 7.2 (3.2) | 10 (1.8) | 0.427 |
| Intensity (lower better) | 1.9 (1.4) | 3.3 (1.0) | 0.421 | 3.7 (1.9) | 4.1 (1.1) | 0.840 |
| Physical (higher better) | 15.3 (2.5) | 15.9 (1.6) | 0.845 | 17.8 (1.6) | 14.4 (1.8) | 0.233 |
| Anxiety 8a (lower better) | 16.5 (4.6) | 12.0 (1.2) | 0.278 | 15.6 (5.3) | 14.2 (2.0) | 0.775 |
| Depression 8a (lower better) | 15.5 (4.0) | 10.7 (0.6) | 0.161 | 13.6 (4.7) | 11.8 (1.3) | 0.638 |
| Self Efficacy 4a (higher better) | 16.3 (1.6) | 15.7 (1.3) | 0.746 | 11.0 (3.5) | 16.4 (1.0) | 0.086 |
| **Cognitive Domains, mean (SD)** |  |  |  |  |  |  |
| Verbal | 24.5 (1.8) | 39.1 (4.3) | **0.031** | 24.0 (2.6) | 36.9 (3.0) | **0.013** |
| Motor | 32.6 (3.6) | 31.6 (2.9) | 0.831 | 33.2 (4.3) | 31.5 (2.3) | 0.701 |
| Spatial | 47.8 (5.8) | 42.8 (5.1) | 0.534 | 50.1 (7.7) | 41.4 (5.4) | 0.367 |
| Processing | 31.8 (2.6) | 40.2 (4.0) | 0.146 | 40.6 (4.9) | 43.2 (4.0) | 0.693 |
| Executive | 40.9 (3.4) | 43.6 (4.8) | 0.677 | 45.0 (4.6) | 48.0 (4.1) | 0.648 |
| Global | 36.3 (2.6) | 39.9 (3.2) | 0.437 | 39.6 (3.3) | 41.4 (3.0) | 0.720 |

## Discussion

Our results suggest that while differences were modest, modified-Mindfulness Based Stress Reduction (m-MBSR) may have some benefit on cognitive function during the early stages of recovery. Though all groups recovered from their baseline visit to follow-up post-intervention without significant differences between them, those undergoing m-MBSR were more likely to improve to a greater extent in more cognitive domains, especially those involving frontal lobes including processing speed and executive function. Interestingly, those in the SSG showed more improvement in their self-reported PROMIS metrics. Importantly, engagement did not necessarily correlate with better results; however, only 2/3 of the participants were able to visibly and actively engage at this point in their recovery.

M-MBSR is a practice thought to alter the brain by engaging the frontal lobes. A meta-analysis of 21 neuroimaging studies in 2014 found that the pre-frontal cortex, anterior cingulate, and orbitofrontal cortex were all involved during mindful meditation, along with the sensory cortex, the insula, and hippocampal regions.^16^ A study of Tibetan monks also showed changes in pre-frontal delta, beta, and gamma activity during deep meditation, with corresponding shifts in proteomics revealing sources located within the attention and emotion networks.^15^ It follows with the hypothesis that activities highly dependent on the frontal lobes, including mood regulation and cognitive tasks dependent on networks involving these regions, may benefit from these changes. Previous studies involving patients with chronic health conditions provide supporting evidence.^17,18,20–30^ However, m-MBSR has not been substantially studied in patients following stroke, and particularly those with minor stroke in the early stages of recovery. In this population, we have demonstrated previously that a lesion in any location can result in difficulties with processing speed and executive function,^13^ and that areas including the bilateral frontoparietal cortices show atypical beta activity,^31^ serving as a potential target for behavioral intervention with MBSR. Clinically, following a course of m-MBSR there does appear to be some benefit, particularly with respect to tasks such as Symbol Digit Modalities written symbol task, and D-KEFS-Trail Making #4, which rely heavily on frontal lobe regions. Studies involving chronic stroke patients showed improvement on these tasks to an even greater degree.^30^ Imaging correlates will need to be further studied.

While cognitive testing results demonstrated small but expected patterns - with patients demonstrating better attention and executive function after m-MBSR compared to those participating in SSG - results for patient-reported outcomes were more surprising. Though both groups had similar PROMIS scores at baseline, SSG patients reported greater improvements in many of the emotional markers following intervention compared to the m-MBSR group. This appears to contradict prior studies of m-MBSR; however, one explanation is that mindfulness involves body awareness and encourages patients to be more in tune with their symptoms. It is possible that following stroke this may have resulted in an increased sensitivity to post-stroke changes and, therefore, increased recognition of the effects of their deficits on their bodies and quality of life. Alternatively, it is possible that the SSG provided greater group cohesion and support – compared to m-MBSR’s focus on more tools-based learning - which might result in inflated subjective reporting of recovery for SSG.

While m-MBSR did appear to result in modest differences in cognition, they were not as large as originally hypothesized. This may be due to several key factors. Groups where brain changes have been observed following mindful meditation, such as Tibetan monks, are typically experienced and actively engaged in the practices.^15^ It would be rational to assume that some level of engagement is required to achieve both clinical and radiographic effects. While patients with minor stroke have low stroke severity overall, there is significant cognitive dysfunction to keep many from returning to work in the subacute setting, potentially posing a challenge to active practice. As the cognitive results did not differ significantly between the intervention groups, evaluating the impact of engagement was necessary. Results did not suggest that level of engagement was a significant predictor of clinical outcome. However, it is critical to point out that when evaluating participants’ levels of involvement in class participation, home practices, and other factors, we found less than 2/3 of participants to be actively engaged in the sessions, and engaged participants appeared, in general, to have better scores at baseline that persisted post-intervention. While the reasoning for the lack of engagement could be simple disinterest in the study, given that individuals voluntarily opted into these sessions, the lack of engagement may indicate that some patients with minor stroke are unable to engage to the level necessary to achieve benefit in the early stages of recovery.

A second possibility for the modest clinical difference lies in the natural history of stroke recovery. As with language and motor deficits, a large majority of cognitive symptoms improve during the initial six months post-infarct.^39^ This makes implementing an intervention during this time attractive to optimize improvement. However, it can also make finding larger differences between groups more difficult, especially when deficits are already less severe at baseline for minor strokes. A more robust difference in results between groups may be expected during the chronic stage of recovery when deficits are more stable. Fortunately, even a small change in the degree of symptoms can make a large difference in a patient’s functional ability following minor stroke. Due to a lack of aphasia or hemiparesis, cognitive impairment is the predominant symptom for many minor stroke survivors, evidenced by scores on the MoCA that are several points lower than expected for age,^37^ resulting in difficulty returning to work.^12^ Conversely, the same small increase in MoCA, as seen following m-MBSR, could significantly improve function.

### Limitations

This study has several limitations. The relatively small cohort of patients with minor stroke was recruited from a single stroke center. In addition, baseline cognitive function was unknown. To account for this, *T*-scores were used to allow for comparison with the general population. Although it is possible that for some individuals a *T*-score <50 was normal, for the entire cohort to have such a poor baseline level given the high average years of education would be unlikely. Additionally, given the multiple administrations of cognitive assessments, it is also possible that some degree of observed improvement may have been due to practice effect. However, alternative versions of tests were used for subsequent evaluation when possible, and tests such as the MoCA have demonstrated reliability even when administered as frequently as every 3 months.^40^ Remote intervention also poses its own unique set of challenges, such as meeting and internet connectivity issues, especially with an older population. The accessibility of taking the class in the comfort of their own home has allowed for greater participation and retention; however, home intervention also includes a greater potential for patients to be distracted. An additional analysis is required to compare the effectiveness of the online version of m-MBSR to the in-person program, as well as determine a formal method for assessing engagement in future studies. Finally, given the clinical nature of this research, the results show correlative rather than causal relationships. Nevertheless, these correlative relationships set the groundwork to continue to find effective interventions for post-stroke patients.

## Conclusions

Despite the aforementioned limitations, this study suggests that m-MBSR may provide a noninvasive, feasible therapeutic approach to improving cognitive deficits following minor stroke, specifically with respect to frontal lobe function. The modest effects seen in this study may be a consequence of m-MBSR requiring a level of engagement that many minor stroke patients in the acute setting are unable to achieve, and therefore m-MBSR may be more beneficial to improving cognition during the chronic stages of recovery; however, even modest effects have the potential to make a significant impact in this group. Future studies are needed to determine if intervention in the subacute setting results in the necessary changes in brain architecture, and whether using a biomarker such as functional neuroimaging to evaluate the integrity of cognitive networks allows for identification of individuals most likely to benefit at various stages post-stroke.

## Data Availability

All data produced in the present study are available upon reasonable request to the authors.

## Acknowledgements

The authors would also like to acknowledge the contributions of all undergraduate members of the Marsh Lab who assisted with data collection for this study.

## References

1. Mozaffarian D, Benjamin EJ, Go AS, et al. Heart Disease and Stroke Statistics—2016 Update. Circulation. 2016;133(4). doi:10.1161/CIR.0000000000000350

2. Feigin VL, Brainin M, Norrving B, et al. World Stroke Organization (WSO): Global Stroke Fact Sheet 2022. International Journal of Stroke. 2022;17(1):18–29. doi:10.1177/17474930211065917

3. Dimyan MA, Cohen LG. Neuroplasticity in the context of motor rehabilitation after stroke. Nat Rev Neurol. 2011;7(2):76–85. doi:10.1038/nrneurol.2010.200

4. Bonita R. Rehabilitation services received by stroke patients: the Auckland stroke study. N Z Med J. 1988;101(854):595–597.

5. Wisenburn B, Mahoney K. A meta-analysis of word-finding treatments for aphasia. Aphasiology. 2009;23(11):1338–1352. doi:10.1080/02687030902732745

6. An M, Shaughnessy M. The Effects of Exercise-Based Rehabilitation on Balance and Gait for Stroke Patients. Journal of Neuroscience Nursing. 2011;43(6):298–307. doi:10.1097/JNN.0b013e318234ea24

7. Brott T, Adams HP, Olinger CP, et al. Measurements of acute cerebral infarction: a clinical examination scale. Stroke. 1989;20(7):864–870. doi:10.1161/01.STR.20.7.864

8. National Institute of Neurological Disorders and Stroke rt-PA Stroke Study Group. Tissue Plasminogen Activator for Acute Ischemic Stroke. New England Journal of Medicine. 1995;333(24):1581–1588. doi:10.1056/NEJM199512143332401

9. Campbell BCV, Mitchell PJ, Kleinig TJ, et al. Endovascular Therapy for Ischemic Stroke with Perfusion-Imaging Selection. New England Journal of Medicine. 2015;372(11):1009–1018. doi:10.1056/NEJMoa1414792

10. Marsh EB, Girgenti S, Llinas EJ, Brunson AO. Outcomes in Patients with Minor Stroke: Diagnosis and Management in the Post-thrombectomy Era. Neurotherapeutics. 2023;20(3):732–743. doi:10.1007/s13311-023-01349-5

11. Makin SDJ, Turpin S, Dennis MS, Wardlaw JM. Cognitive impairment after lacunar stroke: systematic review and meta-analysis of incidence, prevalence and comparison with other stroke subtypes. J Neurol Neurosurg Psychiatry. 2013;84(8):893–900. doi:10.1136/jnnp-2012-303645

12. Marsh EB, Lawrence E, Hillis AE, Chen K, Gottesman RF, Llinas RH. Pre-stroke employment results in better patient-reported outcomes after minor stroke. Clin Neurol Neurosurg. 2018;165:38–42. doi:10.1016/j.clineuro.2017.12.020

13. Marsh EB, Khan S, Llinas RH, Walker KA, Brandt J. Multidomain cognitive dysfunction after minor stroke suggests generalized disruption of cognitive networks. Brain Behav. 2022;12(5). doi:10.1002/brb3.2571

14. Hasenkamp W, Barsalou LW. Effects of meditation experience on functional connectivity of distributed brain networks. Front Hum Neurosci. 2012;(MARCH 2012):1–14. doi:10.3389/fnhum.2012.00038

15. Guo X, Wang M, Wang X, et al. Progressive increase of high-frequency EEG oscillations during meditation is associated with its trait effects on heart rate and proteomics: a study on the Tibetan Buddhist. Cerebral Cortex. 2022;32(18):3865–3877. doi:10.1093/cercor/bhab453

16. Fox KCR, Nijeboer S, Dixon ML, et al. Is meditation associated with altered brain structure? A systematic review and meta-analysis of morphometric neuroimaging in meditation practitioners. Neurosci Biobehav Rev. 2014;43:48–73. doi:10.1016/j.neubiorev.2014.03.016

17. Abbott RA, Whear R, Rodgers LR, et al. Effectiveness of mindfulness-based stress reduction and mindfulness based cognitive therapy in vascular disease: A systematic review and meta-analysis of randomised controlled trials. J Psychosom Res. 2014;76(5):341–351. doi:10.1016/j.jpsychores.2014.02.012

18. de Vibe M, Bjørndal A, Fattah S, Dyrdal GM, Halland E, Tanner-Smith EE. Mindfulness-based stress reduction (MBSR) for improving health, quality of life and social functioning in adults: a systematic review and meta-analysis. Campbell Systematic Reviews. 2017;13(1):1–264. doi:10.4073/csr.2017.11

19. Marsh EB, Girgenti S, Llinas EJ, Brunson AO. Outcomes in Patients with Minor Stroke: Diagnosis and Management in the Post-thrombectomy Era. Neurotherapeutics. 2023;20(3):732–743. doi:10.1007/s13311-023-01349-5

20. Young L, Cappola A, Baime M. Mindfulness Based Stress Reduction: effect on emotional distress in diabetes. Practical Diabetes International. 2009;26(6):222–224. doi:10.1002/pdi.1380

21. DiRenzo D, Crespo-Bosque M, Gould N, Finan P, Nanavati J, Bingham CO. Systematic Review and Meta-analysis: Mindfulness-Based Interventions for Rheumatoid Arthritis. Curr Rheumatol Rep. 2018;20(12):75. doi:10.1007/s11926-018-0787-4

22. Wells RE, Burch R, Paulsen RH, Wayne PM, Houle TT, Loder E. Meditation for Migraines: A Pilot Randomized Controlled Trial. Headache: The Journal of Head and Face Pain. 2014;54(9):1484–1495. doi:10.1111/head.12420

23. He L, Han W, Shi Z. The Effects of Mindfulness-Based Stress Reduction on Negative Self-Representations in Social Anxiety Disorder—A Randomized Wait-List Controlled Trial. Front Psychiatry. 2021;12. doi:10.3389/fpsyt.2021.582333

24. Kabat-Zinn J. An outpatient program in behavioral medicine for chronic pain patients based on the practice of mindfulness meditation: Theoretical considerations and preliminary results. Gen Hosp Psychiatry. 1982;4(1):33–47. doi:10.1016/0163-8343(82)90026-3

25. Kabat-Zinn J, Lipworth L, Burney R. The clinical use of mindfulness meditation for the self-regulation of chronic pain. J Behav Med. 1985;8(2):163–190. doi:10.1007/BF00845519

26. Leung L, Han H, Martin M, Kotecha J. Mindfulness-based stress reduction (MBSR) as sole intervention for non-somatisation chronic non-cancer pain (CNCP): protocol for a systematic review and meta-analysis of randomised controlled trials. BMJ Open. 2015;5(5):e007650. doi:10.1136/bmjopen-2015-007650

27. das Nair R, Cogger H, Worthington E, Lincoln NB. Cognitive rehabilitation for memory deficits after stroke. Cochrane Database of Systematic Reviews. 2016;2016(9). doi:10.1002/14651858.CD002293.pub3

28. Quintana-Hernández DJ, Miró-Barrachina MT, Ibáñez-Fernández IJ, et al. Mindfulness in the Maintenance of Cognitive Capacities in Alzheimer’s Disease: A Randomized Clinical Trial. Journal of Alzheimer’s Disease. 2016;50(1):217–232. doi:10.3233/JAD-143009

29. Paller KA, Creery JD, Florczak SM, et al. Benefits of Mindfulness Training for Patients With Progressive Cognitive Decline and Their Caregivers. American Journal of Alzheimer’s Disease & Other Dementiasr. 2015;30(3):257–267. doi:10.1177/1533317514545377

30. Johansson B, Bjuhr H, Rönnbäck L. Mindfulness-based stress reduction (MBSR) improves long-term mental fatigue after stroke or traumatic brain injury. Brain Inj. 2012;26(13-14):1621–1628. doi:10.3109/02699052.2012.700082

31. Kulasingham JP, Brodbeck C, Khan S, Marsh EB, Simon JZ. Bilaterally Reduced Rolandic Beta Band Activity in Minor Stroke Patients. Front Neurol. 2022;13. doi:10.3389/fneur.2022.819603

32. Homack S, Lee D, Riccio CA. Test Review: Delis-Kaplan Executive Function System. J Clin Exp Neuropsychol. 2005;27(5):599–609. doi:10.1080/13803390490918444

33. Benedict RHB, Schretlen D, Groninger L, Brandt J. Hopkins Verbal Learning Test – Revised: Normative Data and Analysis of Inter-Form and Test-Retest Reliability. Clin Neuropsychol. 1998;12(1):43–55. doi:10.1076/clin.12.1.43.1726

34. Benedict RHB, Schretlen D, Groninger L, Dobraski M, Shpritz B. Revision of the Brief Visuospatial Memory Test: Studies of normal performance, reliability, and validity. Psychol Assess. 1996;8(2):145–153. doi:10.1037/1040-3590.8.2.145

35. Sheridan L, Fitzgerald H, Adams K, et al. Normative Symbol Digit Modalities Test performance in a community-based sample. Archives of Clinical Neuropsychology. 2006;21(1):23–28. doi:10.1016/j.acn.2005.07.003

36. Ashendorf L, Vanderslice-Barr JL, McCaffrey RJ. Motor Tests and Cognition in Healthy Older Adults. Appl Neuropsychol. 2009;16(3):171–176. doi:10.1080/09084280903098562

37. Nasreddine ZS, Phillips NA, Bédirian V, et al. The Montreal Cognitive Assessment, MoCA: A Brief Screening Tool For Mild Cognitive Impairment. J Am Geriatr Soc. 2005;53(4):695–699. doi:10.1111/j.1532-5415.2005.53221.x

38. Marsh EB, Lawrence E, Gottesman RF, Llinas RH. The NIH Stroke Scale Has Limited Utility in Accurate Daily Monitoring of Neurologic Status. Neurohospitalist. 2016;6(3):97–101. doi:10.1177/1941874415619964

39. Girgenti SG, Brunson AO, Marsh EB. Baseline Function and Rehabilitation Are as Important as Stroke Severity as Long-term Predictors of Cognitive Performance Post-stroke. Am J Phys Med Rehabil. 2023;102(2S):S43–S50. doi:10.1097/PHM.0000000000002125

40. Wong A, Yiu S, Nasreddine Z, et al. Validity and reliability of two alternate versions of the Montreal Cognitive Assessment (Hong Kong version) for screening of Mild Neurocognitive Disorder. PLoS One. 2018;13(5):e0196344. doi:10.1371/journal.pone.0196344

